# Kinetics and persistence of the cellular and humoral immune responses to BNT162b2 mRNA vaccine in SARS-CoV-2-naive and - experienced subjects

**DOI:** 10.1101/2022.01.17.22269278

**Authors:** Salomé Desmecht, Aleksandr Tashkeev, Nicole Marechal, Hélène Perée, Yumie Tokunaga, Celine Fombellida Lopez, Barbara Polese, Céline Legrand, Marie Wéry, Myriam Mni, Nicolas Fouillien, Françoise Toussaint, Laurent Gillet, Fabrice Bureau, Laurence Lutteri, Marie-Pierre Hayette, Michel Moutschen, Christelle Meuris, Daniel Desmecht, Souad Rahmouni, Gilles Darcis

**Author notes:** These authors contributed equally to this work (Salomé Desmecht, Aleksandr Tashkeev). These authors also contributed equally to this work (Daniel Desmecht, Souad Rahmouni, Gilles Darcis).

## Abstract

**Background:** Understanding and measuring the individual level of immune protection and its persistence at both humoral and cellular levels after SARS-CoV-2 vaccination is mandatory for the management of the vaccination booster campaign. Our prospective study was designed to assess the immunogenicity of the BNT162b2 mRNA vaccine in triggering the humoral and the cellular immune response in healthcare workers up to 6 months after two doses vaccination.

**Methods:** This prospective study enrolled 208 healthcare workers from the Liège University Hospital (CHU) of Liège in Belgium. All participants received two doses of BioNTech/Pfizer COVID-19 vaccine (BNT162b2). Fifty participants were SARS-CoV-2 experienced (self-reported SARS-CoV-2 infection) and 158 were naïve (no reported SARS-CoV-2 infection) before the vaccination. Blood sampling was performed at the day of the first (T0) and second (T1) vaccine doses administration, then at 2 weeks (T2), 4 weeks (T3) and 6 months (T4) after the 1^st^ vaccine dose administration. A total of 1024 blood samples were collected. All samples were tested for the presence of anti-Spike antibodies using DiaSorin LIAISON SARS-CoV-2 TrimericS IgG assay. Neutralizing antibodies against the SARS-CoV-2 Wuhan-like variant strain were quantified in all samples using a Vero E6 cell-based neutralization-based assay. Cell-mediated immune response was evaluated at T4 on 80 participants by measuring the secretion of IFN-γ on peripheral blood lymphocytes using the QuantiFERON Human IFN-γ SARS-CoV-2, Qiagen. All participants were monitored on weekly-basis for the novo SARS-COV-2 infection for 4 weeks after the 1^st^ vaccine dose administration. We analyzed separately the naïve and experienced participants.

**Findings:** We found that anti-spike antibodies and neutralization capacity levels were significantly higher in SARS-CoV-2 experienced healthcare workers (HCWs) compared to naïve HCWs at all time points analyzed. Cellular immune response was similar in the two groups six months following 2^nd^ dose of the vaccine. Reassuringly, most participants had a detectable cellular immune response to SARS-CoV-2 six months after vaccination. Besides the impact of SARS-CoV-2 infection history on immune response to BNT162b2 mRNA vaccine, we observed a significant negative correlation between age and persistence of humoral response. Cellular immune response was, however, not significantly correlated to age, although a trend towards a negative impact of age was observed.

**Conclusions:** Our data strengthen previous findings demonstrating that immunization through vaccination combined with natural infection is better than 2 vaccine doses immunization or natural infection alone. It may have implications for personalizing mRNA vaccination regimens used to prevent severe COVID-19 and reduce the impact of the pandemic on the healthcare system. More specifically, it may help prioritizing vaccination, including for the deployment of booster doses.

## Introduction

Mass vaccination of the population plays a crucial role in the control of the coronavirus disease 2019 (COVID-19) pandemic. Pioneering studies have shown that the BNT162b2 (Pfizer–BioNTech), as well as the mRNA-1273 (Moderna) vaccines provide strong protective efficacy against COVID-19 in subjects of 16 years old and older (1)(2). Both mRNA vaccines are highly effective in the first few weeks after vaccination against documented infection and symptomatic COVID-19 among adolescents (3)(4). BNT162b2 was proven to be efficient in children younger than 12 years of age (5). As for the mRNA-1273, the clinical trial is still ongoing (Clinical Trial: NCT04796896). Such protection is critical as school-age children may play an important role in the transmission of SARS-CoV-2 (6)(7).

Nevertheless, several studies indicate that immunity gradually waned in all age groups a few months after having received the second dose of vaccine (8)(9)(10)(11). Indeed, 8 months after COVID-19 mRNA vaccination, the median live-virus neutralizing antibody titer, pseudovirus neutralizing antibody titer, and RBD-specific binding antibody titer elicited by the vaccine are significantly lower than the peak titers (12). As a consequence, the rate of confirmed infection among persons vaccinated revealed a substantial increase as the time from vaccination increased (8)(9)(10)(11). Though, COVID-19 mRNA vaccines-induced protection against hospitalization and death persisted with barely any waning for 6 months after the second dose (8)(9)(10)(11), suggesting that persisting cellular immunity drives the immune response and prevents viral dissemination when antibodies disappear. T cell responses persist up to 6 months after vaccination, with the maintenance of a pool of polyfunctional memory antigen-specific T cells (13)(14). mRNA vaccines also produce persisting functional memory B cells (14).

Although the efficacy of the vaccine against severe disease, hospitalization, and death remains high, weakening immunity and emergence of variants of concern create a need for a third vaccine dose (15). Indeed, a very recent study demonstrated that such booster vaccination induces neutralizing immunity even against the new SARS-CoV-2 Omicron harboring 34 mutations more than all the other variants (16). Additional studies has demonstrated a rapid and consistent reduction in the COVID-19 burden among persons living in long-term care facilities after the initiation of a BNT162b2 booster campaign (17) as well as in other groups of age (18). However, information on how pre-existing immunity to SARS-CoV-2 would be boosted by mRNA vaccination remains poorly understood. In particular, the helpfulness and the timing of booster vaccine doses remain to be determined as well as the impact of recovery after SARS-CoV-2 infection. Here, we analyzed the kinetic of humoral response up to 6 months after BNT162b2. We also analyzed cellular response to BNT162b2 6 months after the vaccination. In particular, we studied the impact of previous SARS-CoV-2 infection on immune persistence. All participants were monitored on weekly-based for the novo SARS-CoV-2 infection that may impact immune persistence.

## Methods

### Participant Enrollment and blood sampling

Hospital staff members (including health care workers and administrative staff) of CHU of Liège were invited to participate in the study. Participants were enrolled during February 2021. All participants received two doses of 0.3 mL of BNT162b2 mRNA vaccine administered to the deltoid muscle with a recommended dose interval of 18-21 days. Demographics and clinical data were collected through a questionnaire and included, among others, information regarding previous SARS-CoV-2 infection. All enrolled participants were over 18 years of age. They all acknowledged that they had understood the study protocol and signed the informed consent. Participants were classified into two groups : “experienced group”, that includes participants with reported SARS-CoV-2 infection prior to vaccination, and “naïve group”, consisting of individuals without previous SARS-CoV-2 infection. Blood was collected at the day of the 1st vaccine dose (T0), the day of second vaccine dose (T1), then and at 14 days (T2), one month (T3) and six months (T4) following the second vaccine dose. A total of about 40ml of blood was collected from each subject at each timepoint.

The protocol was approved by the ethics committee (full name: comité d’éthique hospitalo-facultaire universitaire de Liège) of Liège University Hospital (approval number 2021-54).

### Cell mediated immune response to SARS-CoV-2 infection

Cell mediated immune response was assessed by measuring the secretion of IFN-γ by peripheral blood lymphocytes using the QuantiFERON Human SARS-CoV-2 (Qiagen, Cat. No. / ID: 626410). Briefly, blood was collected on four tubes: the control set including one positive and negative one negative tube and the two original Vacutainer tubes containing SARS-CoV-2 antigen 1 (Ag1) and SARS-CoV-2 antigen 2 (Ag2) formulated to activate CD4 T (by Ag1) and both CD4 T and CD8 T (by Ag2) lymphocytes in heparinized whole blood. After blood collection and mixing, tubes were incubated at 37°C for 16 to 24h, IFN-γ was then measured on plasma from the stimulated samples using an CLIA based assay. The threshold for positivity was 0.15 IU/mL as given by Qiagen.

### SARS-CoV-2 detection

Participants were all invited to perform a weekly saliva self-sampling to detect the presence of SARS-CoV-2 following the instruction provided in the starting pack. Participants were asked to return the samples within 12 hours after sampling to the core facility to perform the required follow-up PCR testing. Results were available to them within 24h on the ULiège online platform: https://test-covid.uliege.be/testcovid/index_xt.do.

### Assessment of neutralizing antibodies by sero-neutralization testing (SNT)

SNT analysis were conducted in a specialized biosafety level 3 (BSL3) facility using a SARS-CoV-2 virus Wuhan-like variant (BetaCov/Belgium/Sart-Tilman/2020/1) isolated from a patient hospitalized in March 2020. Virus isolation, expansion, titration and SNT analysis were all performed using nonadherent sub-confluent Vero E6 cells (ATCC® CRL-1586) grown in DMEM supplemented with 2% FBS and penicillin-streptomycin.

The virus stock was titrated in serial log dilutions to obtain a 50% tissue culture infective dose (TCID50) on 96-well culture plates. The plates were observed daily using inverted optical microscope for five days to evaluate the presence of cytopathic effect (CPE) and the end-point titer was calculated according to the Reed & Muench method based on 2 × 3 replicates.

Serum test samples were heat-inactivated for 40 min at 56 °C and two-fold serial dilutions, starting from 1:10 up to 1:320, were performed in triplicate in DMEM/FBS on 96-well culture plates. Sera dilutions (50 μl/well) were then mixed with an equal volume of a pre-titrated viral solution containing 100 TCID50 of SARS-CoV-2 virus. The serum-virus mixture was incubated 1 h at 37 °C in a humidified atmosphere with 5% CO2. After incubation, 100 μl of a Vero cells suspension were added so that 20,000 cells were deposited in each well (19). The plates were then re-incubated for 5 days. For each serum, the process was repeated twice by two different, trained people. After 5 days, CPE was evaluated under light microscopy by two independent persons. Serum dilutions showing CPE were considered as non-neutralizing (negative), while those showing no CPE were considered neutralizing/positive. Virus sero-neutralization titer was reported as the highest dilution of serum that neutralizes CPE in 50% of the wells (NT50). If results from the 2 duplicate plates were discordant, these samples were processed again in a subsequent SNT session. For all sera showing a NT50>1:320, a second process was made using higher dilutions (up to 1:20,480). Positive (NT50=1:160, from the Belgian National Reference Centre) and negative (saline) controls were inserted in each plate.

### Assessment of total anti-spike IgGs

The DiaSorin LIAISON SARS-CoV-2 TrimericS IgG assay (DiaSorin, Stillwater, USA), a chemiluminescent immunoassay using magnetic particles coated with recombinant trimeric SARS-CoV-2 spike protein, was used for quantitative determination of IgG antibodies in human serum samples. The assay was performed on LIAISON XL analyzer (DiaSorin) according to the manufacturer’s instructions.

### Data analysis

All the analyses were carried out in R version 4.1.1. All codes and data to reproduce the results are available at : https://github.com/tashkeev-alex/vaccination_study.

### Estimating the decay rates

Decay rates of anti-Spike IgG and NT50 were estimated as previously reported (14)(20)(21), i.e. by using linear mixed model with censoring, implemented in *lmec* R package (22). Considering the sampling timeline in this study, we preferred to model the dependency of the response variables on the number of days post-vaccine by using single-slope rate using the data starting from T2. Namely, the model is:

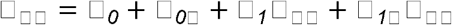

where □_□□_ is log of anti-Spike IgG or negative log of NT50 for individual *i* at time point *j* post-vaccine 2^nd^ dose, *β*_*0*_ is global intercept, □_*0*□_ is a patient-specific adjustment to *β*_*0*_, *β*_*1*_ is a fixed slope dedicated to capture the average decay rate across individuals, □_□□_ is number of days post-vaccine 2^nd^ dose for individual *i* at time point *j*, and □_*1*□_ is a patient-specific adjustment to *β*_*1*_. Likelihood ratio test was used to detect whether rates were different between naive and SARS-CoV-2 experienced individuals.

### Separating individuals with neutralizing versus non-neutralizing antibodies by anti-S IgG

Logistic regression of neutralizing status at the last time point (T4 at 6 months post-vaccine, 1^st^ dose) on the log of anti-Spike IgG was used to determine the threshold immunoglobulin value that provides the best separation of the classes. Sensitivity and specificity of the model were estimated by summarizing those across five stratified folds of cross-validation (no hyperparameter tuning or model selection was performed).

### Testing for differences or associations among immune and clinical parameters

In all corresponding procedures we used non-parametric statistics, i.e. either tie-corrected Spearman correlation coefficient or Wilcoxon rank-sum test. We used log-transformed anti-Spike IgG, NT50, and IFN-γ values. In multiple linear regression models, we centered and scaled the continuous predictors by two standard deviations allowing the comparison of coefficients for continuous and binary predictors on the same scale (23). In case of continuous dependent variables, we centered and scaled them in the same way. In case of multivariate dependent variables (anti-S IgG or NT50 across time points), we centered and scaled them across individuals within each time point.

## Results

### Cohort characteristics, demographics and samples collection

Our study included 208 consenting subjects among the CHU of Liège staff members who received the Pfizer–BioNTech BNT162b2 vaccine during February 2021. Among them, 158 were SARS-CoV-2 naive subjects while 50 had COVID-19 prior to vaccination (experienced SARS-CoV-2). The cohort was skewed toward females (∼80%). The age was however balanced between naive (34-53 years old) and experienced groups (37-55 years old) with median of 43 and 46 respectively. The BMI was also balanced between the two groups with a median of 24,8 (21,3 to 27,7) for naïve and 24,0 (21,4 to 27,3) for experienced. Comorbidities of the participants included asthma (11% in naive group and 6,2% in experienced group), autoimmunity autoimmunity (5,3% in naive group - 3 with Hashimoto’s thyroiditis, 3 with hypothyroidism, 2 with Chron’s disease, 1 with psoriasis, 1 with diabetes and rheumatoid arthritis, and 2,1% in experienced group - 2 with Lupus and 1 with psoriatic arthritis), blood cancer (non-active, 2% in naive group and 0% in experienced group), other cancers (non-active, 3,9% in naive group and 4,2% in experienced group) and immunodeficiency (1,3% in naive group and 2,1% in experienced group (**Table 1**).

**Table 1.** Cohort description.

| **Characteristic** | **naive, N = 158<sup>†</sup>** | **recovered, N = 50<sup>†</sup>** |
| --- | --- | --- |
| **Sex** |  |  |
| F | 125 (80%) | 39 (80%) |
| M | 31 (20%) | 10 (20%) |
| Unknown | 2 | 1 |
| **Age** |  |  |
|  | 43 (34, 53) | 46 (37, 55) |
| Unknown | 2 | 1 |
| **Smoking** |  |  |
|  | 25 (16%) | 5 (10%) |
| Unknown | 2 | 1 |
| **BMI** |  |  |
|  | 24.8 (21.3, 27.7) | 24.0 (21.4, 27.3) |
| Unknown | 8 | 2 |
| **Asthma** |  |  |
|  | 17 (11%) | 3 (6.2%) |
| Unknown | 5 | 2 |
| **Autoimmunity** |  |  |
|  | 8 (5.3%) | 1 (2.1%) |
| Unknown | 6 | 2 |
| **Immunodeficiency** |  |  |
|  | 2 (1.3%) | 1 (2.1%) |
| Unknown | 7 | 2 |
| **Blood Cancer** |  |  |
|  | 3 (2.0%) | 0 (0%) |
| Unknown | 5 | 2 |
| **Other Cancer** |  |  |
|  | 6 (3.9%) | 2 (4.2%) |
| Unknown | 5 | 2 |
| **# time points** |  |  |
| 3 | 9 (5.7%) | 4 (8.0%) |
| 4 | 33 (21%) | 5 (10%) |
| 5 | 116 (73%) | 41 (82%) |
| <sup>†</sup>n (%); Median (IQR) |  |  |

In total, 1024 samples were collected. Sampling was performed the day of 1^st^ and 2^nd^ dose of the vaccine (T0 and T1), then 2 weeks, 4 weeks and 6 months after the 1^st^ dose (T2, T3 and T4). 73% of naive participants donated blood at all time points while 21% and 5,7% donated blood 4 and 3 times respectively. In the experienced group, 82% of subjects donated blood at all time points while 10% and 8% donated 4 and 3 times respectively (**Table 1**).

### Dynamics of humoral response to SARS-CoV-2 mRNA vaccine BNT162b2

We assessed the level of circulating trimeric Spike IgG in plasma samples using DiaSorin LIAISON SARS-CoV-2 TrimericS IgG assay. The level of anti-Spike antibodies clearly discriminates naive and experienced groups at T0 (day of the 1^st^ vaccine dose). Median across experienced subjects was equal to 91.5 BAU/ml (4.8-772) confirming pre-exposition to the virus. The level of anti-Spike antibodies increased rapidly after the 1^st^ vaccine dose in both groups but with higher titer in the experienced group over the naive one (497 BAU/ml in naive, 6630 BAU/ml in SARS-CoV-2 experienced). The anti-Spike circulating antibodies levels reached their maximum two weeks after the 2^nd^ vaccine dose and start declining two weeks later. This decrease continued over time between two and six months after the 1^st^ vaccine dose with a half-life of 55 days. The kinetics of anti-Spike antibodies production was similar between the naive and experienced groups. The levels of produced antibodies were however significantly higher in the experienced group over the naive group at all time points (**Figure 1 A, B**).

**Figure 1.**
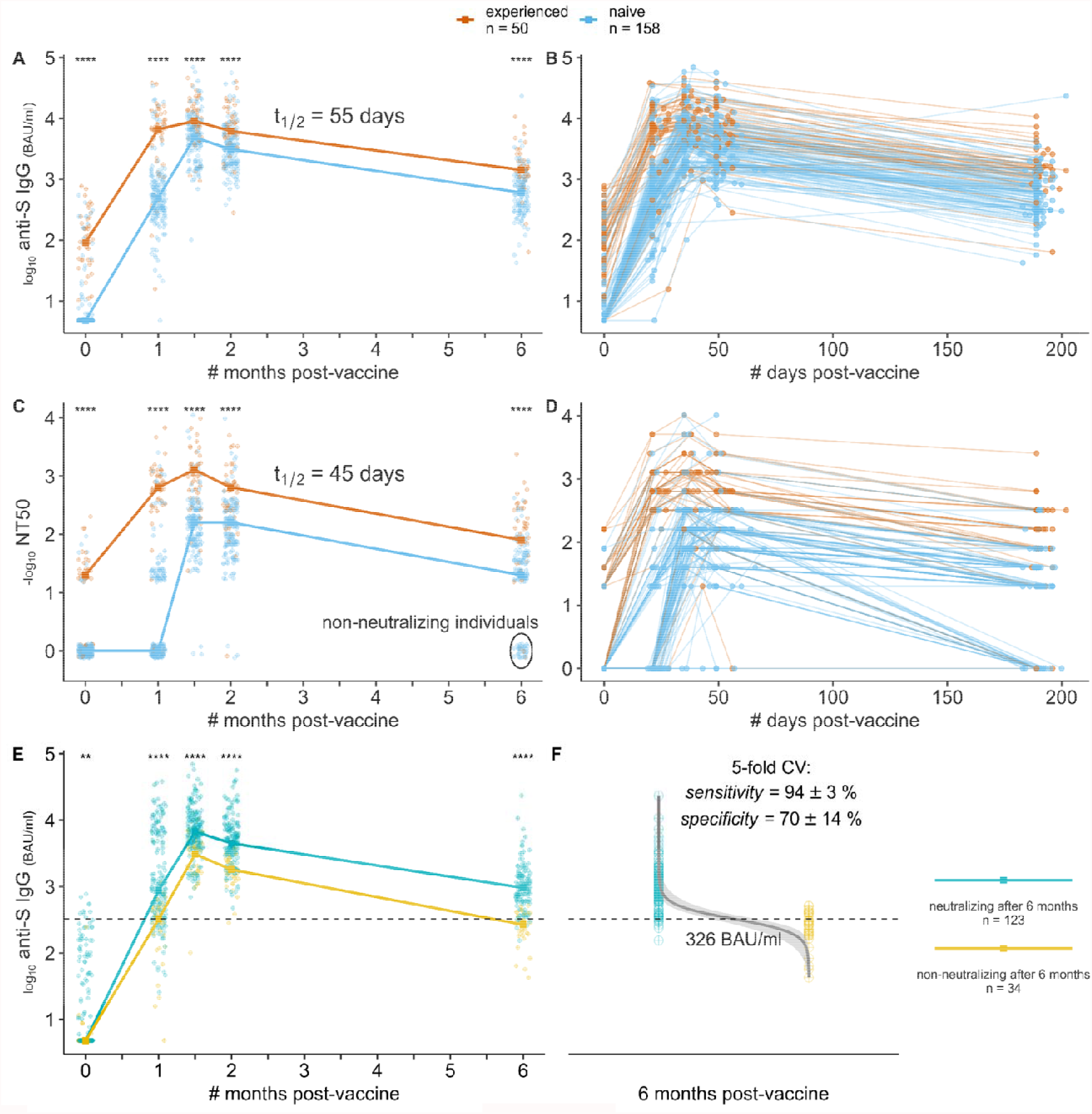
Dynamics of humoral immune response to SARS-CoV-2 mRNA vaccine BNT162b2. **(A-B)** dynamics of anti-Spike IgG *(log10 BAU/ml)* – median across individuals per approximate time point (**A**), or for each individual (**B**) with exact time points; **(C-D)** dynamics of virus neutralization titer *(-log10 values, zeroed if titer was above 1/20)* – median across individuals per approximate time point (C), or for each individual with exact time points (D); **(E)** same as in (**A**), but grouping individuals by the ability to neutralize virus at the last time point *(based on (****C***)); **(F)** Logistic regression-derived boundary value of anti-Spike IgG level at the last time point separating individuals with no detectable neutralization at the last time point from the others *(inability to separate them by anti-Spike IgG at earlier time points is shown on S1 Fig)*, mean ± standard deviation values of the model performance are shown.

To evaluate the quality of secreted antibodies, we measured neutralization capacities at all time points and all subjects using previously reported protocols (19). More than a half of SARS-CoV-2-experienced participants (26/50) showed a detectable neutralization (mostly 1:20) at the day of the vaccine 1^st^ dose administration (T0) in contrast to subjects with no prior SARS-CoV-2 infection (naive group) (only 9/158) (**Figure 1C, D**). Some of them with detectable neutralization could be either undiagnosed SARS-CoV-2 experienced individuals, or show cross-reactivity due to previous infections by other coronaviruses. One month after the 1^st^ vaccine dose (T1), median neutralization titer increased substantially to 1:640 in experienced group, while no such an increase was observed in the vast majority (only in 63/158) of the naive group. Two weeks after the 2^nd^ vaccine dose (T3), almost everyone developed a neutralizing ability, disregarding the previous diagnosed viral exposure. Despite the SARS-CoV-2 experienced participants showing higher neutralization capacity at all time points including those after full vaccination, the decay rate was similar in both participants types, namely the half-life period of 45 days after the 2^nd^ dose (**Figure 1C, D**). Noteworthy, the neutralization capacity at the last time point (6 months after the 1^st^ vaccine dose) has decreased to non-detectable level in some individuals, mostly in those who also showed poor response to the 1^st^ vaccine dose (26/31 of naive and 3/3 of SARS-CoV-2 experienced) (**Figure 1C, D**).

In clinical practice, measuring antibodies neutralization capacities is complicated and time consuming. Therefore, and since measuring anti-Spike IgG level is routinely used, we modeled the ability to use anti-Spike antibodies levels to predict whether a person still has some neutralization capacity at some reasonably far time point after vaccination (6 months in the case of our study) (**Figure 1E, F)**. Logistic regression classifier allowed to separate individuals with no detectable neutralization at the last time point and the others by the anti-Spike IgG at the last time point (sensitivity: mean=94%, std=3%; specificity: mean=70%, std=14% across five stratified folds of cross-validation). The decision value estimated on the whole dataset was 326 BAU/ml. Using anti-Spike IgG level at earlier time points did not allow the classification (**Figure S1**).

### Association of BNT162b2-induced anti-SARS-CoV-2 neutralizing antibodies with participants’ clinical parameters

Because of the variable level of the anti-Spike circulating antibodies’ quantity and quality between vaccinated (naive and experienced) subject, we assessed the association of total antibodies as well as neutralizing status of individuals at the last time point with a reasonable set of available clinical parameters such as sex, smoking status, age, and body mass index (BMI) in a single model containing also one of the parameters representing the anti-Spike IgG level (**Figure 2**). By the latter we considered SARS-CoV-2 infection history based on participant’s self-reporting (**Figure 2A**), anti-Spike IgG level before vaccination (T0) (**Figure 2B**), after 2 doses vaccination (T2) (**Figure 2C**), or 6 months after vaccination (T4) (**Figure 2D**). Among all tested clinical parameters, only age was significantly associated with the neutralizing status (decreasing chances to have neutralizing capacity), and even its effect size was far less (1.4-5.3x) than that of the parameter representing anti-Spike IgG level. No effect of sex, smoking status, and BMI were detected at any significance level (**Figure 2**).

**Figure 2.**
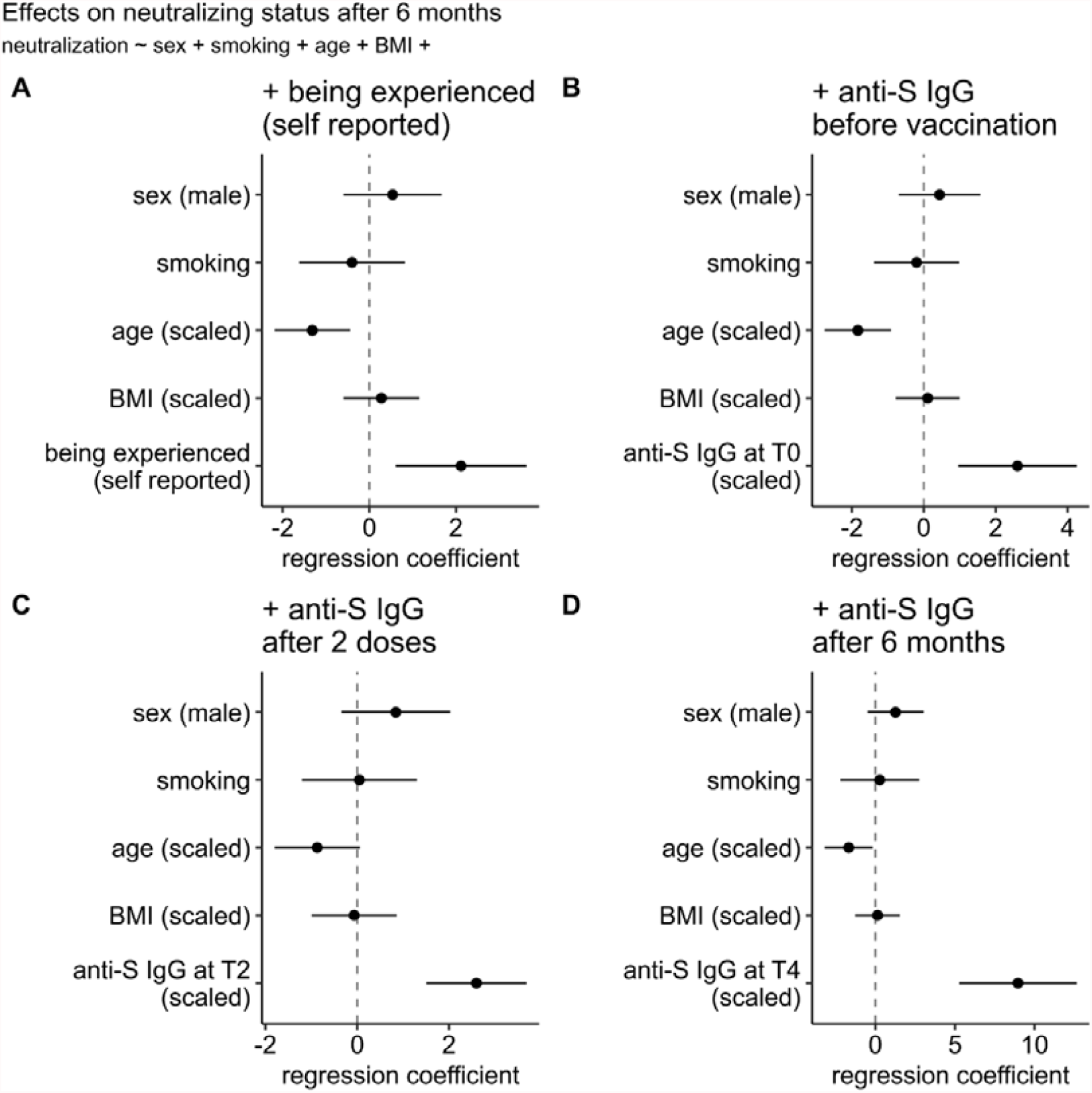
Effects of clinical parameters on neutralizing status at six months after 1^st^ vaccination dose. given the (A) experience status, (B) anti-Spike IgG level at 1^st^ time point *(T0* : *day of the 1*^*st*^ *vaccine dose)*, (C) anti-Spike IgG level at T2 time point *(after 2*^*nd*^ *vaccine dose)*, or (D) anti-Spike IgG level at T4 *(6 months after vaccination)*. Regression coefficients for all predictors can be compared on the same scale since continuous predictors were centered and scaled by two standard deviations

We next tested the association of humoral response parameters themselves with the abovementioned set of clinical parameters across all time points (**Figure 3**). Since NT50 and anti-Spike IgG levels were strongly correlated at each time point for both SARS-CoV-2-experienced and naive individuals (**Figure 3A**), we tested them in separate models using either multivariate (**Figure 3B, D**) or univariate (**Figure 3C, E**) multiple linear regression, i.e. considering values across time points as a single vector or separately. Again, only age showed a significant association with the outcomes explaining 25% and 21% of conditional variance in multivariate models for anti-Spike IgG level and NT50, respectively. Interestingly, effects of age in univariate models were negative (i.e. decreases humoral response parameter value) at all time points except the first one, at which the effect was significant but of the opposite sign. One plausible hypothesis could be that older individuals are more likely to face cold coronaviruses before, and thus are more likely to show some baseline humoral response to SARS-CoV-2.

**Figure 3.**
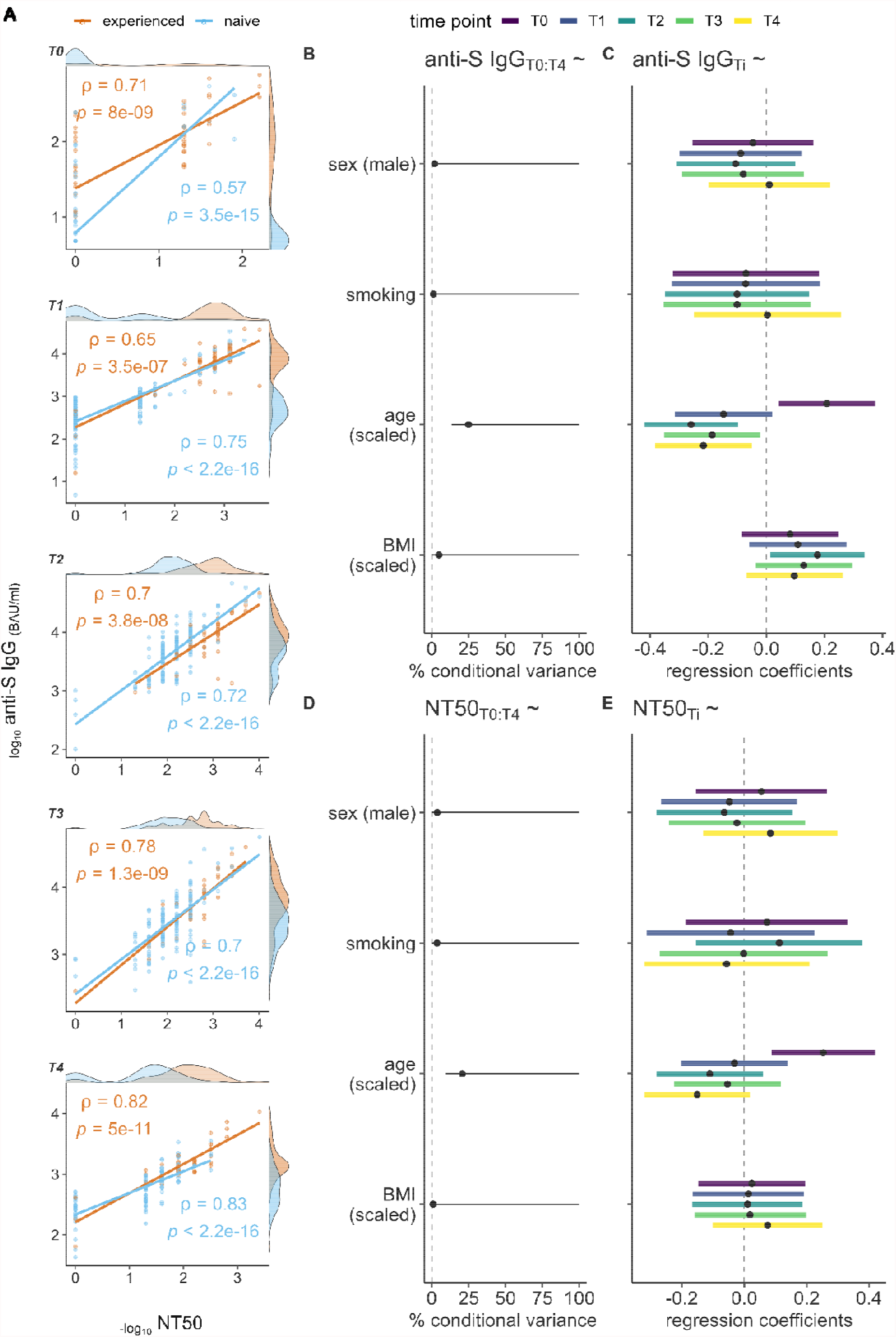
Associations between humoral immune response and clinical parameters across time points. **(A)** Spearman correlation (rho) between neutralization titer and level of anti-Spike IgG for each time point *(T1-T5)*; **(B)** Effects of clinical parameters on level of anti-Spike IgG across time points in a multivariate multiple regression model *(left)*, or in a univariate multiple regression model per time point *(right)*; **(C)** Same as (B) but for NT50 instead of anti-Spike IgG level.

### Cellular immune response to SARS-CoV-2 mRNA vaccine BNT162b2

To evaluate T-cell specific immune response to BNT162b2 vaccine, heparinized whole blood was stimulated with SARS-CoV-2 specific peptides contained in the QuantiFERON-SARS-CoV-2 tubes designed to activate both CD4T and CD8T cells. IFN-γ was then measured by CLIA. Since this assay was only commercially available after we completed the collection of samples at T0 to T3, we only assayed cellular immune response at the last time point (T4) (**Figure 4**). Cellular response was evaluated on 80 participants with 63 in the naive group and 17 in the experienced group. In terms of IFN-γ secretion, we did not observe any significant difference between naive and experienced groups for none of the antigen used (Ag1 and Ag2) (**Figure 4A, B**). However, when subjects were clustered based on their neutralizing capacities at 6 months after vaccination, the difference was highly significant between those with high neutralizing titers versus the one with no detectable neutralization (**Figure 4B**). We next investigated if there was any association between IFN-γ secretion, anti-Spike total antibodies and neutralizing antibodies titers. As shown in figure 4A, there was no correlation between anti-S IgG and IFN-γ in the naive group for both Ag1 and Ag2. Interestingly, IFN-γ secretion was highly correlated to anti-Spike IgG in the experienced group (**Figure 4C top panel**). IFN-γ secretion was also correlated to neutralization titers in all groups, although the correlation was most significant in the experienced group for Ag1 (p=3e-4) over the naive group (p=0,0011) and vice versa for Ag2 (p= 0.0026 for experienced and p=0,00029 for naive) (**Figure 4C lower panel**). There was no statistically significant association between cellular immune response parameters and any of the available clinical parameters (**Figure S3**).

**Figure 4.**
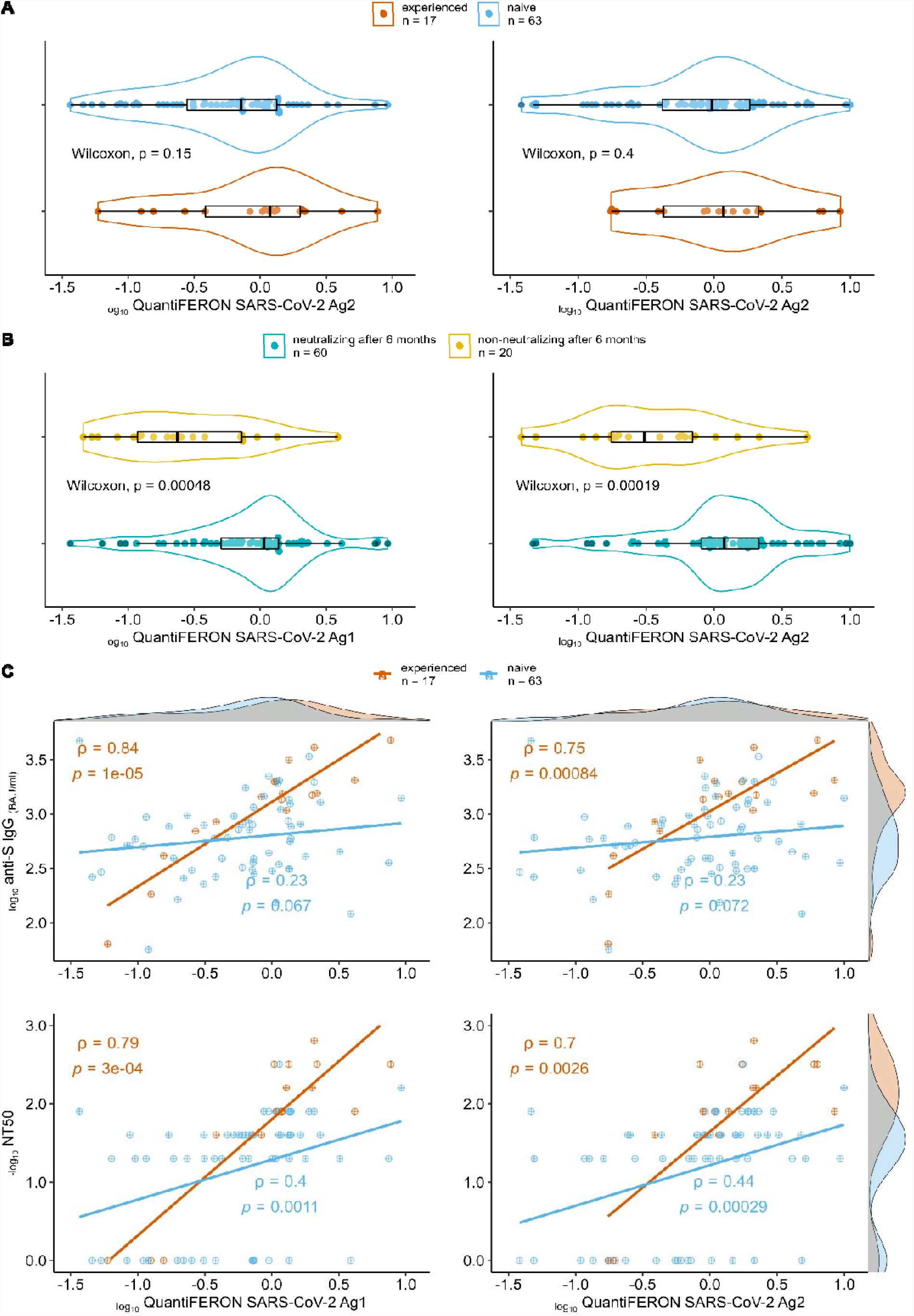
Associations between cellular and humoral responses in naïve versus SARS-CoV-2 experienced groups. (**A**) Comparison of cellular immune response (QuantiFERON-SARS-CoV-2 Ag1 (left panel) and QuantiFERON-SARS-CoV-2 Ag2 (right panel) between SARS-CoV-2 experienced (orange) and naïve (blue) groups. (**B**) Same as (A) but comparing individuals with no detectable neutralization at the 6 months after 1^st^ vaccine dose (yellow) and those with neutralizing titers (blue) at the same time point. (**C**) Spearman correlation (rho) between anti-Spike IgG level (top panels) or neutralization titer (bottom panels) with cellular response represented by QuantiFERON-SARS-CoV-2 Ag1 (left) or Ag2 (right).

### COVID-19 testing of the study participants

Participants were all invited to perform a weekly saliva self-sampling to detect the presence of SARS-CoV-2. Nevertheless, the adherence to the weekly SARS-CoV-2 sampling rapidly dropped down **(Figure S4)**. Only one participant was tested as positive during the study timeline.

## Discussion

Our study confirms previous evidence for an earlier, stronger and more persistent humoral immune response in individuals previously infected with SARS-CoV-2 versus naïve individuals following vaccination with the BNT162b2 mRNA vaccine (24). The anti-spike antibody and neutralization capacity levels six months after vaccination protocol were significantly higher in SARS-CoV-2 experienced HCWs compared to naïve HCWs. We did not observe such differences regarding cellular immune response six months following 2^nd^ dose vaccination. Reassuringly, most participants had a detectable cellular immune response to SARS-CoV-2 six months after vaccination. These findings are in line with a recent observation from Samanovic and colleagues who also reported more pronounced differences in humoral, over cellular responses between individuals SARS-CoV-2-naive versus recently SARS-CoV-2 experienced subjects (25).

Besides the impact of SARS-CoV-2 infection history on immune response to BNT162b2 mRNA vaccine, we observed a significant association between age and persistence of humoral response. The more elderly is a participant, the less durable was the humoral response. This is in line with results from other studies demonstrating a similar decrease of anti-SARS-CoV-2 antibodies in all age groups few months after the second vaccine dose, especially among 65 years-old or older persons (26). Regarding cellular immune response, although not statistically significant, we observed a trend towards a negative impact of age.

Our results confirm previous findings indicating that anti-spike antibodies are very well correlated with neutralizing antibodies (27)(28). This may allow to set a threshold of anti-spike antibodies predicting neutralization capacity with a high sensitivity. Although correlates of protection from SARS-CoV-2 are not fully defined yet, Khoury and colleagues showed that neutralization level is highly predictive of immune protection, reinforcing the results of other reports suggesting that neutralization titer is an important predictor of vaccine efficacy (21)(29)(30). Our findings may thus have important implications for waning vaccine immunity and vaccination strategies. Our results may also contribute to better define patients who are at high risk of developing severe COVID-19 and who could benefit from anti-SARS-CoV-2 mAb products (PMID: 35016195) (PMID: 34087172). Available stocks of mAbs that retained activity against Omicron are extremely limited in most settings, while many vulnerable patients can be considered as eligible for this treatment. Predictors of remaining neutralization capacity may help clinicians with the difficult selection of patients for whom this intervention would be most beneficial.

The humoral immune response as measured through quantification of anti-spike IgG or neutralizing antibodies correlated with cellular immune responses six months following vaccination. Interestingly, this correlation was much stronger in the group of participants with a history of SARS-CoV-2 infection, indicating that the level of anti-spike antibodies in this group may not only predict the level of neutralizing antibody but also the level of cellular immunity as well.

In conclusion, our data strongly reinforce the relevance of previous SARS-CoV-2 infection for understanding vaccine immune responses. It may have implications for personalizing mRNA vaccination regimens used to prevent severe COVID-19 and reduce the impact of the pandemic on the healthcare system. More specifically, it may help prioritizing vaccination, including for the deployment of booster doses.

## Data Availability

All data produced in the present work are contained in the manuscript

## Author Contributions

“Conceptualization: DD, SR, GD; methodology: DD, SR, GD; formal analysis: SD, AT, DD, SR, GD; investigation: SD, AT, NM, HP, YT, LL, CF, BP, CL, MW, MM, NF, LL, MPH; writing-original draft preparation, SD, AT, DD, SR, GD; review and editing: all authors; supervision: LG, FB, SR, MPH, MM, CM, GD, DD; funding acquisition: GD, SR, MM. All authors have read and agreed to the published version of the manuscript.”

## Institutional Review Board Statement

The study was conducted according to the guidelines of the Declaration of Helsinki, and approved by the Ethical Committee (comité d’éthique hospitalo-facultaire universitaire de Liège, approval number 2021-54).

## Informed Consent Statement

Informed consent was obtained from all subjects involved in the study

## Data Availability Statement

Data is contained within the article or supplementary material

## Acknowledgments

We are very thankful to all HCW participants in this study. We are also very thankful to Jean-Baptiste Giot, Frederic Frippiat, Anne-Sophie Sauvage, Philippe Leonard, Raphaël Schils, Sébastien Bontemps, Cecile Meex and Margaux Dandoy for clinical assistance and scientific helpful discussions.

## Supplementary information

### Supplementary figures

**Figure S1:**
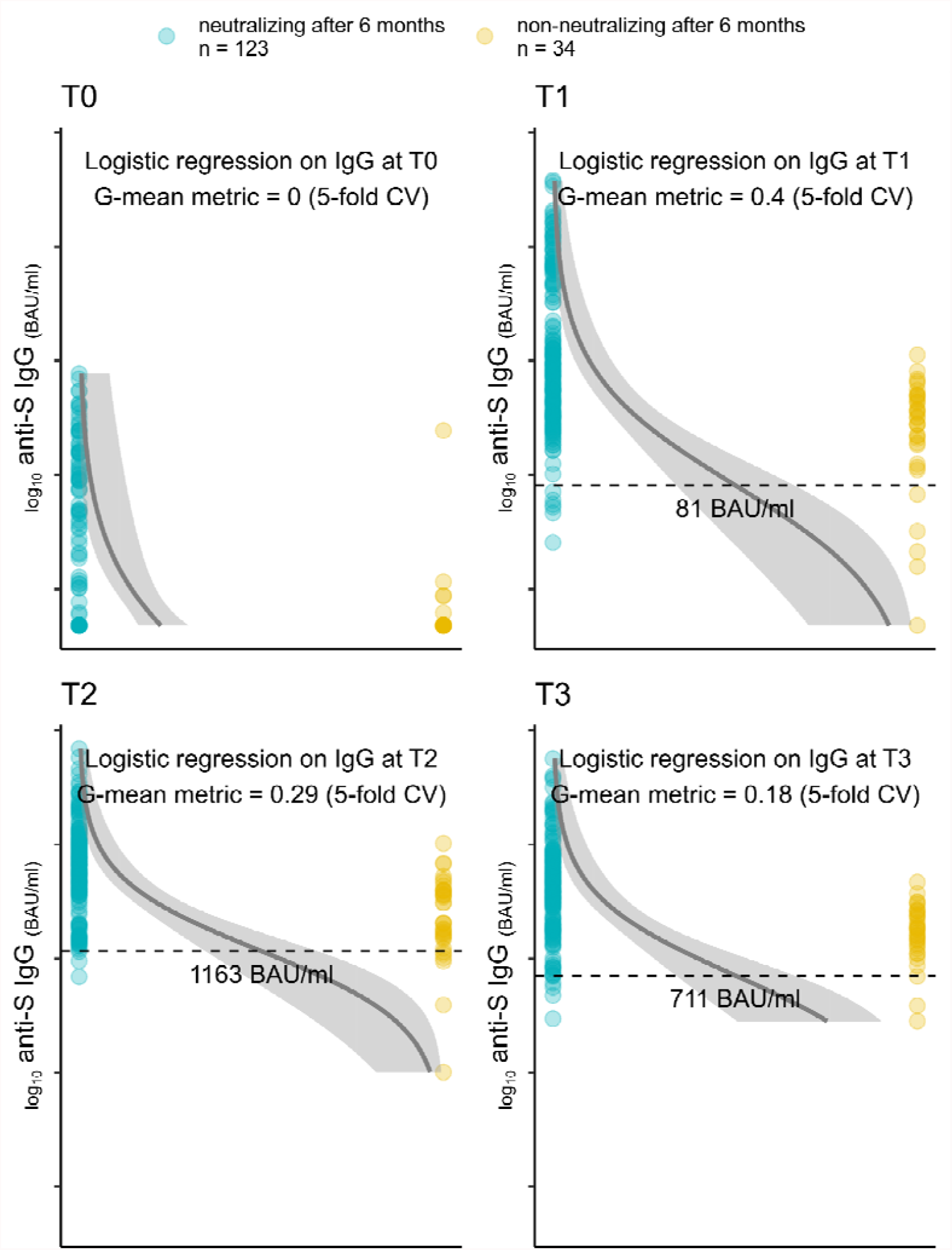
Inability to separate individuals neutralizing and non-neutralizing at last time point by anti-Spike IgG values at earlier time points. G-mean is a geometric mean of sensitivity and specificity, a single metric used to evaluate the model.

**Figure S2:**
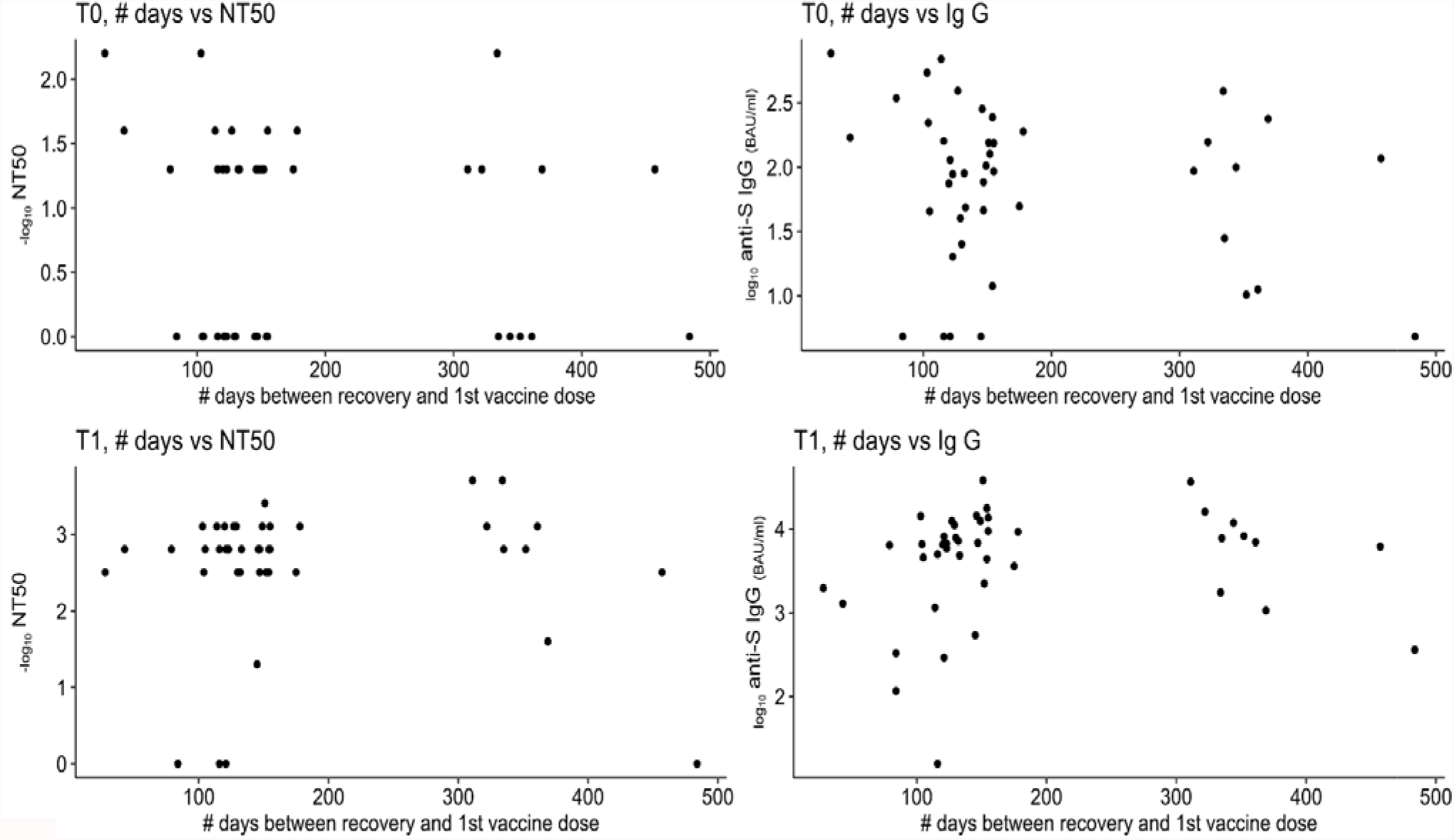
Absence of impact of time after recovery on humoral response parameters before and after 1st vaccine dose.

**Figure S3:**
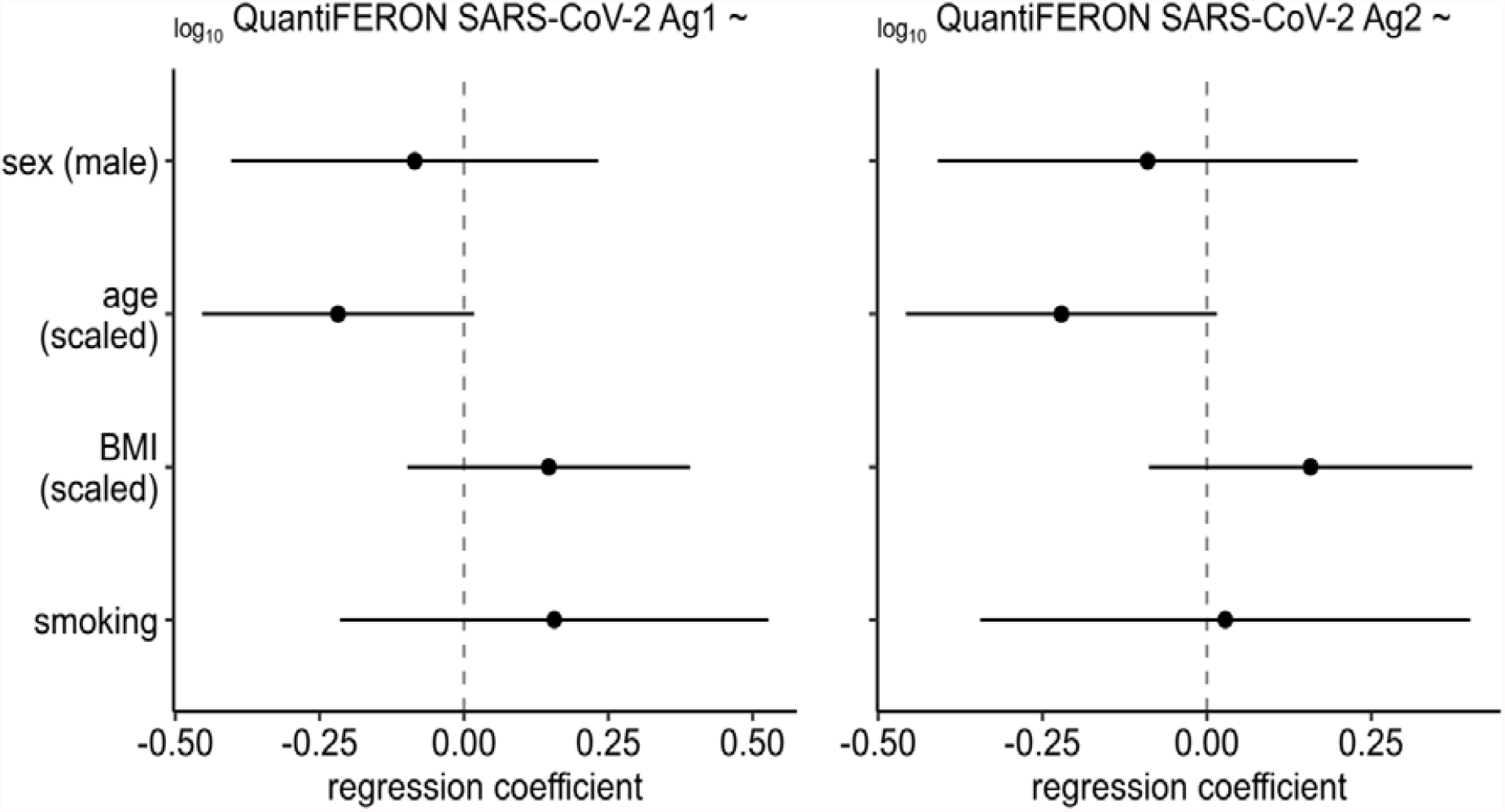
Effects of clinical parameters on cellular response at the last time point.

**Figure S4:**
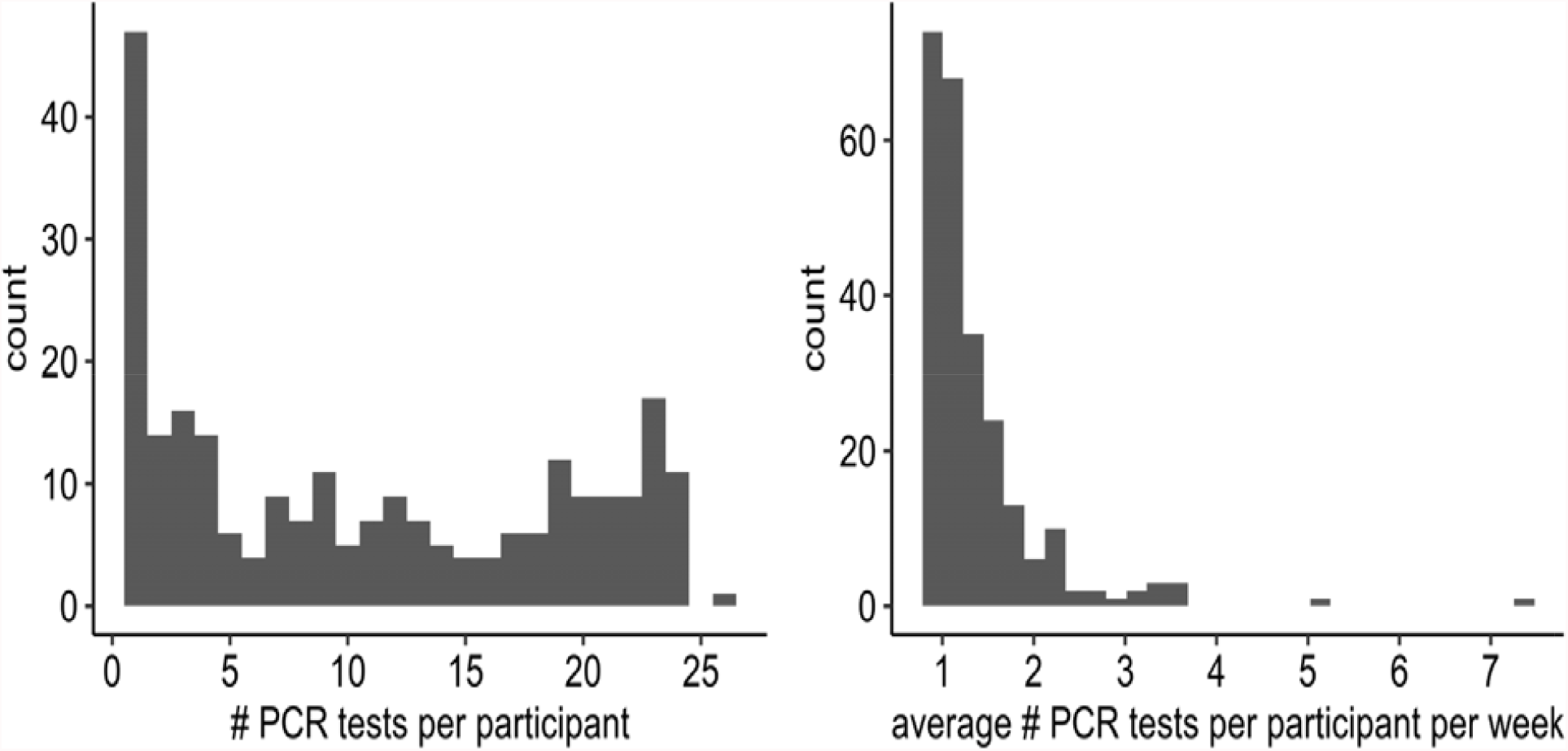
Statistics of COVID-19 testing of the study participants. Only one participant was tested as positive during the study timeline.

